# Clinically Accessible Phenotypes Refine Cardiovascular Risk Stratification Beyond Contemporary Clinical Equations

**DOI:** 10.64898/2026.09.10.26362697

**Authors:** Hongxi Yang, Ju Guo, Xinxin Shi, Kejia Ma, Yuhan Jiang, Dingpeng Li, Xin Zhou, Ying Yu, Dandan Huang

**Author notes:** Corresponding authors: **Address for correspondence:** Ying Yu, PhD, Department of Pharmacology, Tianjin Medical University, 22 Qixiangtai Road, Tianjin 300070, China; and Dandan Huang, PhD, Department of Pharmacology, Tianjin Medical University, 22 Qixiangtai Road, Tianjin 300070, China. These authors contributed equally (Hongxi Yang, Ju Guo, Xinxin Shi, Kejia Ma).

## Abstract

**BACKGROUND:** Contemporary cardiovascular risk equations guide prevention, yet cardiovascular risk remains distributed beyond the highest-risk groups. Whether broader baseline phenotyping can improve risk stratification and whether this information can be reduced to clinically accessible measures is uncertain.

**OBJECTIVES:** To assess whether baseline phenotyping improves cardiovascular risk stratification beyond PREVENT and CHARGE-AF and whether this information can be compressed into clinically accessible measures.

**METHODS:** We studied 266,410 UK Biobank participants without prevalent CHD, HF, AF, or stroke, divided into training, validation, and held-out test samples. PREVENT equations were used to estimate 10-year risk for CHD, HF, and stroke, and CHARGE-AF to estimate 5-year AF risk. Models considered 3,838 baseline phenotypes. Geographic replication was assessed in Scotland and Wales.

**RESULTS:** The highest-risk 20% identified by the clinical equations accounted for 44.8% to 55.2% of subsequent events across the 4 outcomes. Broad phenotyping refined risk stratification beyond these equations. After compression from 3,838 baseline phenotypes, disease-specific sets of 13 to 19 clinically accessible measures retained part of the incremental discrimination provided by broader phenotyping, yielding held-out IPCW AUC improvements of 0.013 to 0.030 relative to the corresponding clinical equations, with the largest gain for HF. At the same 20% prioritization level, the clinically accessible measures increased the proportion of subsequent events captured by 2.7 to 4.9 percentage points. Findings were directionally consistent in geographic replication.

**CONCLUSIONS:** Disease-specific sets of clinically accessible measures retained part of the information identified from broad phenotyping and refined cardiovascular risk stratification beyond contemporary clinical equations. Independent external validation is required before clinical use.

**CONDENSED ABSTRACT:** Among 266,410 UK Biobank participants, the highest-risk 20% defined by PREVENT for CHD, HF, and stroke and CHARGE-AF for AF accounted for 44.8% to 55.2% of subsequent events.

Broad baseline phenotyping refined risk stratification, and part of this incremental prognostic information was retained after compression from 3,838 baseline phenotypes to disease-specific sets of 13 to 19 clinically accessible measures. Findings were directionally reproduced across UK Biobank regions, but independent external validation is required before clinical use.

## INTRODUCTION

Cardiovascular risk equations help clinicians match preventive assessment and treatment to a patient’s likelihood of future cardiovascular events.^1–3^ PREVENT and CHARGE-AF estimate disease-specific risk from a deliberately small set of established clinical predictors.^4–6^ Their performance varies across populations,^7,8^ and even well-calibrated equations cannot concentrate every future event within the fraction of a population that a health system can assess most intensively.

Several information layers can refine conventional risk estimates. Polygenic risk scores (PRSs) capture inherited susceptibility,^9–11^ whereas social and environmental measures describe risk context not represented in most equations.^12–14^ Circulating biomarkers, proteomic and metabolomic profiles, and electrocardiographic artificial intelligence also provide incremental prognostic information.^15–20^ Recent studies have begun to combine these layers or reduce high-dimensional measurements to sparse signatures.^21–27^ Most, however, have focused on one cardiovascular outcome or one measurement modality. Whether the same baseline information is useful across major cardiovascular diseases, and how much of that information can be retained in clinically accessible measures, remains unresolved.

We examined this question across coronary heart disease (CHD), heart failure (HF), atrial fibrillation (AF), and stroke using a broad set of 3,838 phenotypes measured at recruitment. We quantified the proportion of future events captured within the highest-risk 20% defined by disease-specific clinical equations and assessed whether broad baseline phenotyping refined risk stratification beyond these equations. We then determined how much of this incremental information could be retained by reducing the broad phenotype set to parsimonious disease-specific sets of clinically accessible measures. We also examined whether these clinically accessible measures provided additional prognostic information after accounting for disease-specific polygenic risk and whether the findings were consistent across UK Biobank regions.

## METHODS

### Study population and outcomes

UK Biobank recruited more than 500,000 adults aged 40 to 69 years between 2006 and 2010.^28^ We included 266,410 unrelated participants of genetically inferred White British ancestry with quality-controlled genotype data, prospective follow-up, and no prevalent coronary heart disease (CHD), heart failure (HF), atrial fibrillation (AF), or stroke at baseline. Participants were randomly assigned in a 50:20:30 ratio to training (n = 133,205), validation (n = 53,282), and held-out test (n = 79,923) samples. Training data were used for model estimation, validation data for model selection, and the held-out sample for final evaluation. Incident CHD, HF, AF, and stroke were identified from linked hospital admission and mortality records through December 19, 2022. Prediction horizons were 10 years for CHD, HF, and stroke and 5 years for AF. The study was conducted under UK Biobank Application 83974. UK Biobank received ethical approval from the North West Multi-centre Research Ethics Committee (REC reference 11/NW/0382; extended 21/NW/0157), and all participants provided written informed consent.

### Clinical comparators and assessment levels

Clinical comparators were the published component-specific PREVENT equations for CHD and stroke, the 10-year PREVENT-HF base equation, and CHARGE-AF for 5-year AF risk.^4–6^ PREVENT-HF was calculated without optional predictors using preventr version 0.12.0. Published coefficients were used for the primary analyses, with training-sample recalibration examined separately as a sensitivity analysis. Predictor mapping and implementation details are provided in the **Supplemental Methods** and **Supplemental Table 1**. Because the 4 disease-specific equations do not share a common treatment threshold, risk stratification was compared across prespecified fractions of the population prioritized for further assessment (10%, 15%, 20%, 25%, and 30%), with 20% used as the principal reference for presentation and the full 10% to 30% range used to evaluate robustness to the choice of prioritization fraction. Risk-score cutoffs were defined in the validation sample and applied unchanged in the held-out sample.

### Baseline phenotypes and model development

Among 7,226 prespecified candidate variables, 3,838 unique phenotypes measured at recruitment met predefined timing and data-quality criteria. Phenotypes were grouped into 6 phenotypic domains and 6 measurement modalities (**Supplemental Table 2**). All eligible phenotypes were considered irrespective of clinical accessibility; restrictions based on routine availability or assessment burden were introduced only when deriving clinically accessible measure sets. Disease-specific polygenic risk scores (PRSs) were generated using PRS-CS.^29^ For each outcome, Cox models incorporated the clinical-equation linear predictor as an offset. Candidate phenotype models were fitted in the training sample and selected according to validation-set IPCW AUC. Coefficients were re-estimated in the combined training and validation samples before final evaluation in the held-out sample. Elastic-net regression was used for multivariable phenotype selection.^30^ The held-out sample was not used for model or threshold selection. We evaluated whether baseline phenotypes provided prognostic information beyond the corresponding clinical equation and whether this information remained informative after accounting for the disease-specific PRS. Additional details on preprocessing, missing data, phenotype coding, and PRS construction are provided in the **Supplemental Methods**.

### Event prioritization and supporting analyses

At each assessment level, a future event was considered newly included when the participant was outside the group prioritized by the clinical equation but inside the equally sized group prioritized after addition of phenotypic information. An event was considered displaced when the reverse occurred. The net difference between newly included and displaced events was expressed as the change in the proportion of future events captured at the same prioritization level. This change was evaluated across the prespecified 10% to 30% range, with the 20% level used for the principal comparison in the main figures. Shapley decomposition was used to estimate the contribution of individual phenotypic domain-by-modality groups to the change in future events captured at the 20% prioritization level, and overlap between contributing groups was assessed at the 20% level. As supporting analyses, phenotype-outcome associations contributing more strongly to risk stratification were compared with those contributing little or none with respect to association strength, split-sample replication, genetic support, and recurrence across cardiovascular outcomes.^31–36^ These analyses were used for interpretation and did not influence phenotype selection.

### Clinically accessible measure sets

Clinical accessibility was defined independently of associations with study outcomes. Candidate measures were restricted to routinely recorded measurements and brief additional assessments, including self-reported, symptom, or functional assessments and selected additional blood or urine assays, while specialized measurements and variables duplicating predictors already included in the clinical equations were excluded. Disease-specific sets were restricted to no more than 20 clinically accessible measures. Validation data were used to select the smallest sets that retained as much of the incremental discrimination of the clinically accessible reference model as feasible while maintaining a positive average improvement in future-event capture across the 10% to 30% prioritization range. When the prespecified retention target could not be achieved within this limit, the best-performing capped set was selected in the validation sample. Full selection criteria are provided in the Supplemental Methods. We also examined whether similar sets of measures were selected after accounting for the disease-specific PRS.

### Geographic replication

For geographic replication, model development was repeated among participants from England using separate training (n = 175,632) and validation (n = 58,710) samples. The resulting models were then evaluated in participants from Scotland and Wales without further fitting or recalibration. The published disease-specific clinical equation, without PRS, served as the comparator. Selected phenotypes, coefficients, numbers of clinically accessible measures, and risk-score cutoffs were determined entirely within the England development samples before geographic evaluation.

### Statistical analysis

Discrimination was assessed at the prespecified prediction horizon using IPCW AUC^37^ and Harrell C-index,^38^ and calibration using the calibration slope and observed-to-expected event ratio.^39^ Ninety-five percent confidence intervals were estimated from 1,000 paired participant-level bootstrap samples. All statistical tests were 2-sided. Additional statistical procedures and software versions are provided in the **Supplemental Methods**.

### Data sharing Statement

Individual-level UK Biobank data are available to approved researchers through the UK Biobank Access Management System, subject to the relevant application and data-use requirements. Aggregate data underlying the principal figures are provided with the submission. Analysis and figure-generation code, together with synthetic test inputs, is publicly available at https://github.com/Code-ui-2607/CVD_EWAS. No individual-level UK Biobank data are included in the repository.

## RESULTS

The study included 266,410 participants, of whom 133,205 were assigned to the training sample, 53,282 to the validation sample, and 79,923 to the held-out test sample (**Supplemental Figure 1A**). In the disease-specific held-out samples, there were 1,286 CHD events among 75,089 participants, 1,022 HF events among 77,900 participants, 1,105 AF events among 75,367 participants, and 961 stroke events among 75,089 participants (**Supplemental Table 3**; **Supplemental Figures 1B and 1C**). In the held-out sample, IPCW AUCs for the published clinical equations were 0.755 for CHD, 0.772 for HF, 0.746 for AF, and 0.720 for stroke (**Supplemental Figure 2A**). Recalibration using the training sample improved agreement between predicted and observed risk but, as expected, did not materially alter discrimination (**Supplemental Figure 2B**).

**Figure 1.**
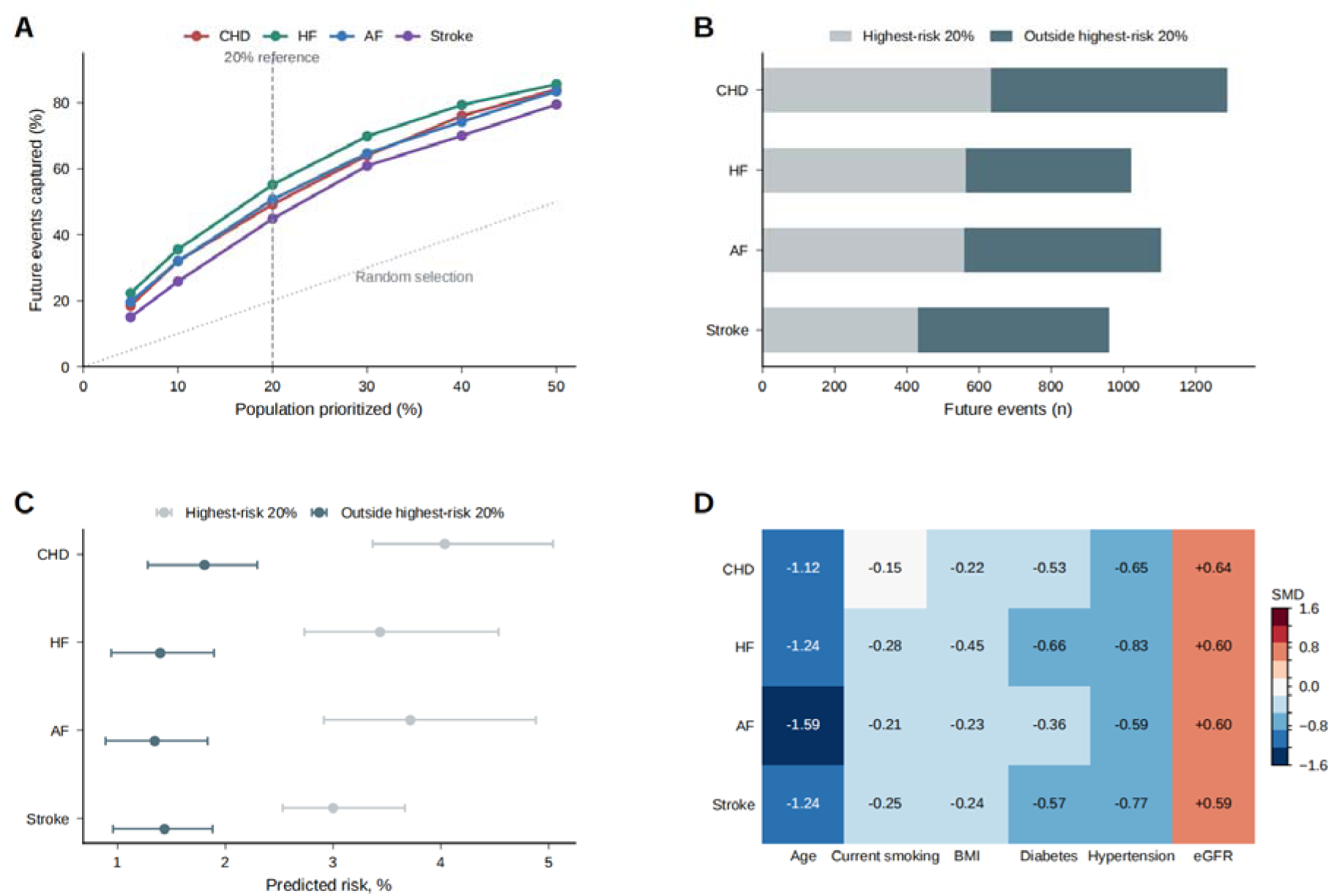
Cardiovascular Events Across Clinical Risk Strata **A**, Future CHD, HF, AF, and stroke events captured as the proportion prioritized increased from 5% to 50%; the vertical line marks the 20% reference, and the diagonal line indicates random prioritization. **B**, Events within and outside the highest-risk 20%. **C**, Predicted risk among future cases within and outside the highest-risk 20%; points are medians and lines indicate interquartile ranges. **D**, Standardized mean differences in baseline characteristics, calculated as outside minus within the highest-risk 20%. AF indicates atrial fibrillation; BMI, body mass index; CHD, coronary heart disease; eGFR, estimated glomerular filtration rate; HF, heart failure; SMD, standardized mean difference.

### Clinical risk stratification and event distribution

Although the clinical equations concentrated subsequent cardiovascular events among individuals at higher predicted risk, risk remained distributed across the population (**Figure 1A**). Across the 4 outcomes, the highest-risk 20% accounted for 44.8% to 55.2% of subsequent events (**Figure 1B**). Predicted risk was lower among future cases outside this group than among cases within the highest-risk group (**Figure 1C**). Compared with future cases in the highest-risk group, those occurring outside the group were younger, had lower body mass index and higher estimated glomerular filtration rate, and were less likely to smoke or have diabetes or hypertension (**Figure 1D**). Thus, the clinical equations concentrated approximately half of subsequent events within the 20% of participants at highest predicted risk, whereas future cases outside this group generally had less prominent conventional risk profiles.

### Incremental value of baseline phenotyping

Baseline phenotypes spanned all 6 prespecified phenotypic domains and 6 measurement modalities (**Figure 2A**). The final broader phenotype models contained 38 to 553 phenotypes across outcomes. When added to the corresponding clinical equations, broad phenotyping improved held-out discrimination across all four outcomes (ΔIPCW AUC, 0.015 to 0.041; **Supplemental Table 5**). At the 20% prioritization level, broad phenotyping increased the proportion of future events captured by 4.0 to 8.0 percentage points across outcomes (**Figure 2B**). This reflected 9.3 to 14.0 percentage points of newly included events, partly offset by 4.4 to 6.5 percentage points of displaced events.

**Figure 2.**
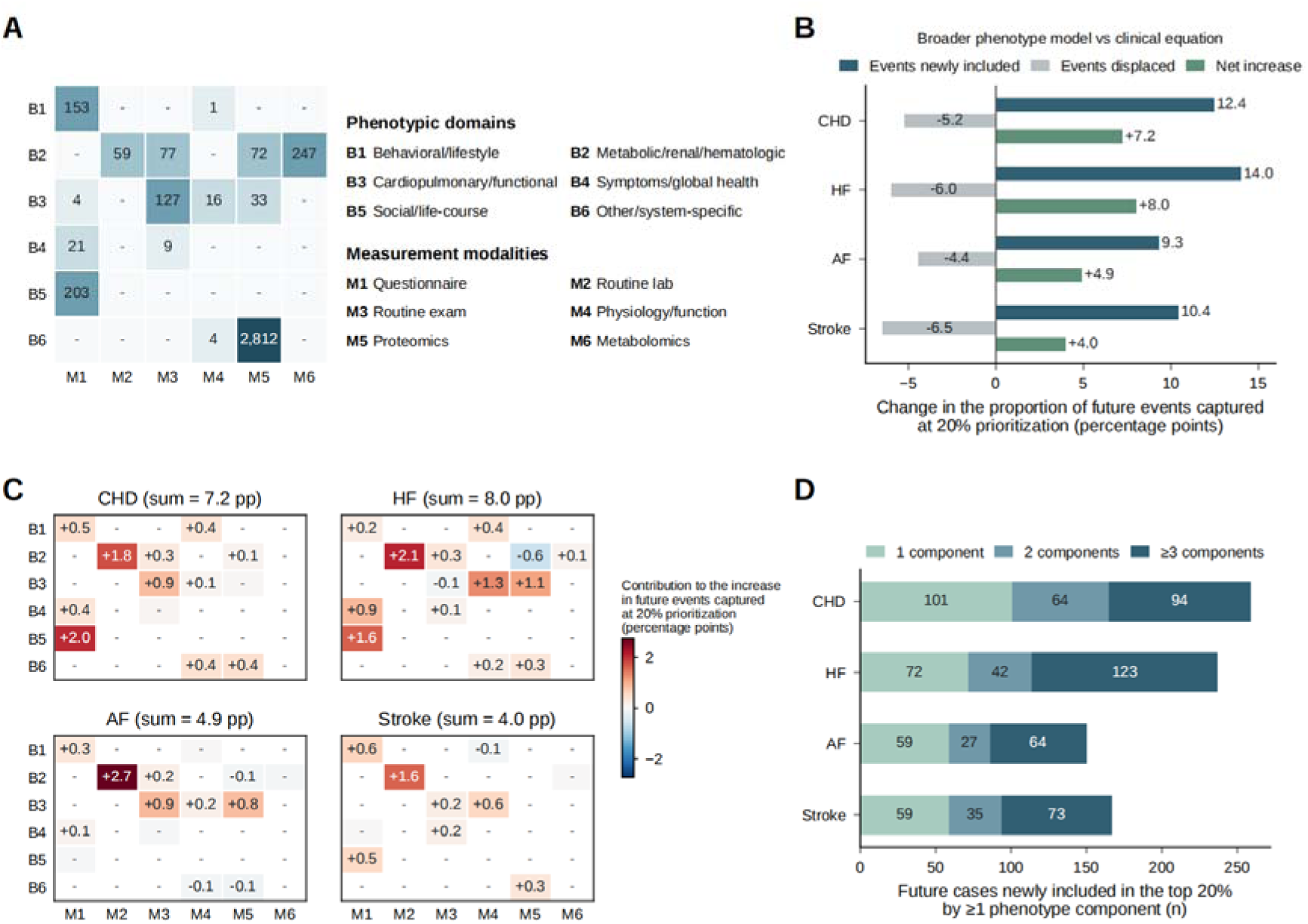
Incremental Value of Baseline Phenotyping **A**, Distribution of 3,838 baseline phenotypes across 6 phenotypic domains and 6 measurement modalities. **B**, Events newly included, displaced, and the net increase in the proportion of future events captured for the broader phenotype model versus the clinical equation at 20% prioritization. Newly included events were within the group prioritized by the phenotype model but not the clinical equation; displaced events were defined conversely. **C**, Shapley decomposition of the increase in future events captured at 20% prioritization across domain-by-modality components. **D**, Future cases newly included in the top 20% by at least 1 phenotype component, grouped by the number of contributing components. B1-B6 indicate behavioral/lifestyle, metabolic/renal/hematologic, cardiopulmonary/functional, symptoms/global health, social/life-course, and other/system-specific domains; M1-M6 indicate questionnaire/self-report, routine laboratory, routine examination, physiology/function, proteomics, and metabolomics.

The incremental information arose from multiple phenotypic domains and measurement modalities rather than a single source (**Figure 2C**). At the 20% prioritization level, 150 to 259 future cases were newly included by at least 1 phenotype component across outcomes, and approximately 61% to 70% were included by 2 or more components (**Figure 2D**). Incomplete overlap and only partial correlation between component scores further supported complementary contributions across phenotypic domains and measurement modalities (**Supplemental Figures 3A to 3D**).

Phenotype-outcome associations contributing most strongly to risk stratification were more frequently supported by several complementary analyses, including statistical association, split-sample replication, cross-outcome recurrence, and genetic support from LDSC and Mendelian randomization (**Supplemental Figures 4A and 4B**). The likelihood of high contribution increased progressively as associations recurred across a greater number of cardiovascular outcomes (**Supplemental Figure 4C**). Validation-defined high-contribution associations were also substantially more likely to retain a positive contribution in the held-out sample than associations with lower or no validation contribution (pooled OR: 32.57; 95% CI: 20.67-51.32) (**Supplemental Figure 4D**). Together, these analyses showed that high-contribution associations tended to recur across cardiovascular outcomes and remain informative in the held-out sample.

### Risk refinement with clinically accessible measures

We next evaluated whether the incremental information from broad phenotyping could be captured with a smaller set of measures suitable for routine clinical assessment. From the 3,838 baseline phenotypes, disease-specific sets of 13 to 19 clinically accessible measures were selected (**Figure 3A**). The selected measures differed across outcomes, although several measures of general health and hematologic status were shared across diseases (**Figure 3B**). The complete composition of these measure sets is provided in **Supplemental Table 4**.

**Figure 3.**
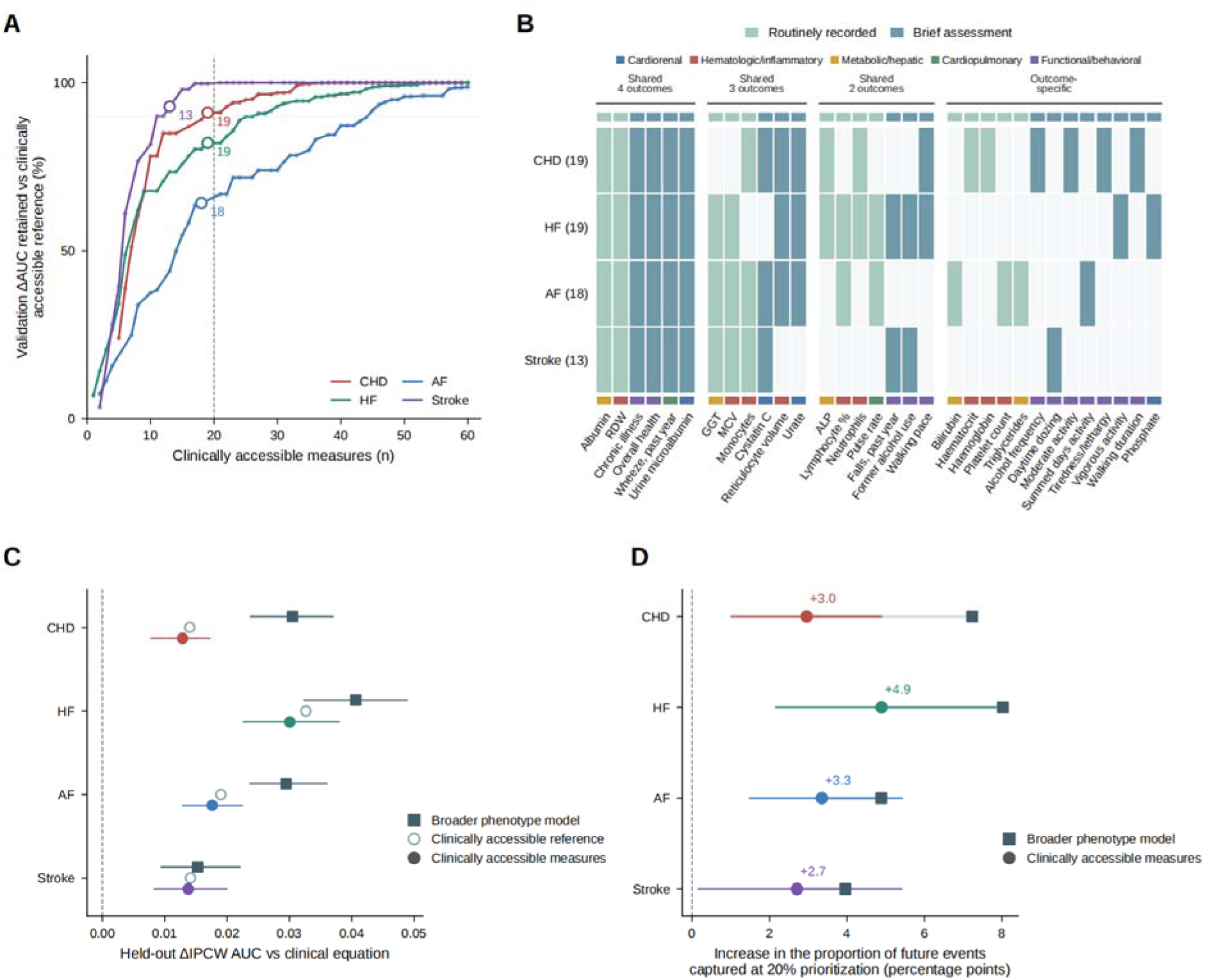
Clinically Accessible Measures **A**, Validation AUC retention as clinically accessible measures increased; reference lines mark the 90% target and 20-measure limit, and labels indicate the final number of measures for each outcome. **B**, Final disease-specific measure sets grouped by the number of outcomes sharing each measure. The upper strip denotes acquisition tier, which defined clinical accessibility; the lower strip shows the clinical dimension assigned after model selection for interpretation and did not enter model fitting. **C**, Held-out increment in horizon-specific IPCW AUC over the clinical equation for the broader phenotype model, clinically accessible reference model, and final clinically accessible measure set. **D**, Increase in the proportion of future events captured at 20% prioritization for the broader phenotype model and final clinically accessible measures. Error bars indicate 95% CIs. AF indicates atrial fibrillation; AUC, area under the receiver operating characteristic curve; CHD, coronary heart disease; HF, heart failure; IPCW, inverse probability of censoring weighting.

When added to the corresponding clinical equations, the clinically accessible measures increased held-out IPCW AUC by 0.013 to 0.030 across outcomes, with the largest improvement for HF (**Figure 3C; Supplemental Table 5**). These smaller sets retained 42% to 90% of the discrimination gain from broader phenotyping, depending on the outcome. At the 20% prioritization level, they increased the proportion of future events captured by 2.7 to 4.9 percentage points across outcomes (**Figure 3D**).

We further examined whether the phenotypic information was complementary to inherited genetic risk. The incremental contribution of disease-specific PRS varied substantially across outcomes, being more pronounced for CHD and AF and more modest for HF and stroke. In contrast, clinically accessible measures provided additional discrimination and increased the proportion of future events captured across all four outcomes even after accounting for PRS (**Supplemental Figures 5A and 5B**). Conversely, PRS retained additional prognostic information beyond the clinically accessible measures for some outcomes, supporting overall complementarity between phenotypic and genetic risk information. Measure sets derived with and without PRS adjustment were highly similar (**Supplemental Figure 5C**) and achieved nearly identical discrimination when evaluated in addition to the clinical equation and PRS (**Supplemental Figure 5D**). Thus, although the direct incremental value of PRS differed across diseases, conditioning phenotype selection on PRS had little effect on the selected measures or their predictive performance. Corresponding performance estimates are provided in **Supplemental Table 6**.

### Geographic replication

To assess geographic reproducibility, models developed in England were evaluated in Scotland and Wales without refitting or recalibration (**Figure 4A**). The evaluation included 21,692 to 31,195 participants and 293 to 551 events across outcomes. As in the main held-out analysis, the highest-risk 20% accounted for a substantial proportion of subsequent events (**Figure 4B**).

**Figure 4.**
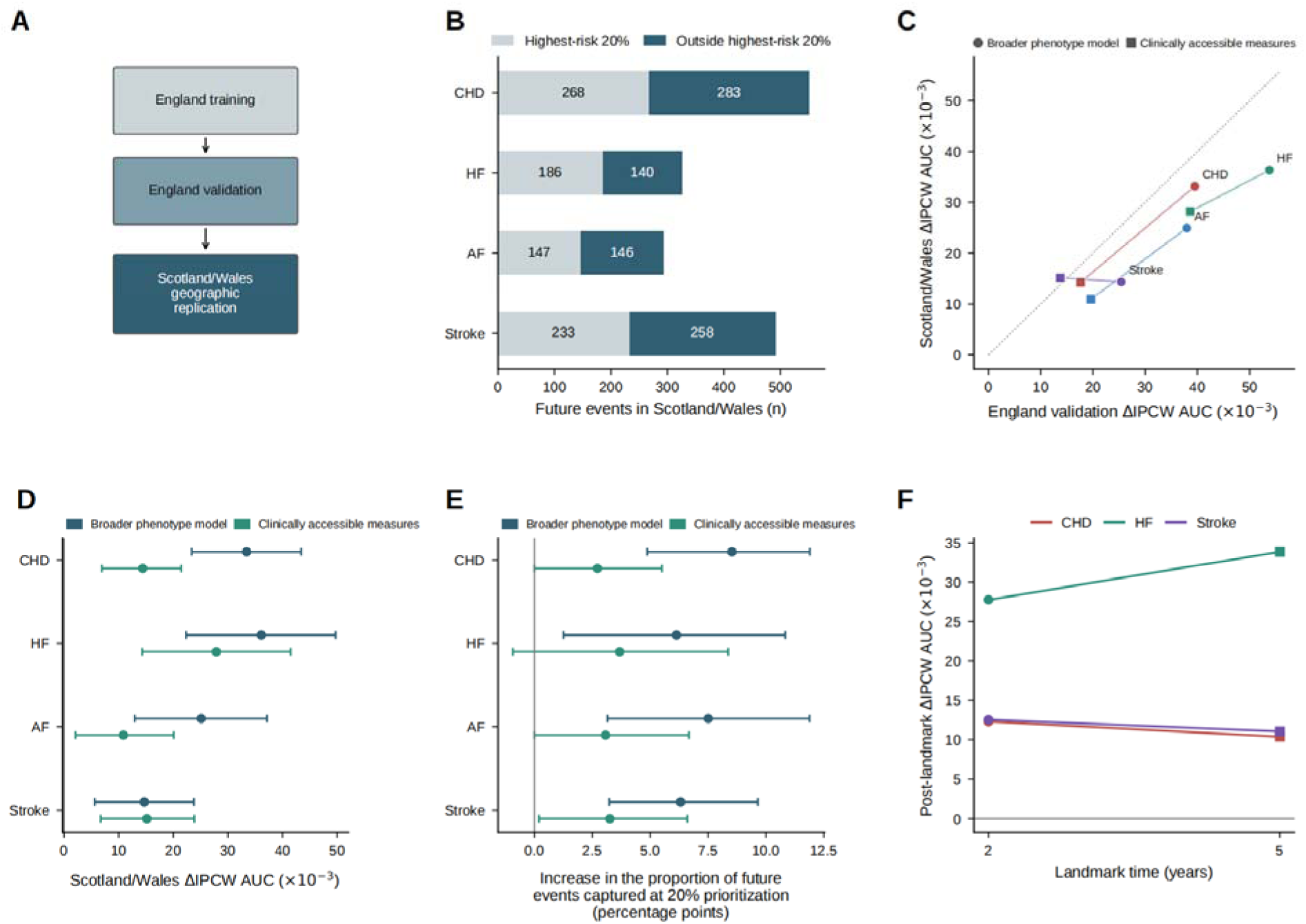
Geographic Replication in Scotland and Wales **A**, Development and validation in England followed by evaluation in Scotland and Wales without refitting or recalibration. **B**, Future events within and outside the highest-risk 20% in Scotland and Wales. **C**, Incremental IPCW AUCs in England validation versus Scotland/Wales evaluation; the diagonal line indicates identity. **D**, IPCW AUC increment for the England-derived broader phenotype model and clinically accessible measures. **E**, Increase in the proportion of future events captured at 20% prioritization for the broader phenotype model and clinically accessible measures. **F**, IPCW AUC increment after excluding events within 2 or 5 years for CHD, HF, and stroke; AF was omitted because its horizon was 5 years. Error bars indicate 95% CIs. AF indicates atrial fibrillation; AUC, area under the receiver operating characteristic curve; CHD, coronary heart disease; HF, heart failure; IPCW, inverse probability of censoring weighting.

Incremental discrimination was directionally concordant between England validation and Scotland/Wales evaluation (**Figure 4C**). In Scotland and Wales, the broader phenotype models increased IPCW AUC by 0.015 to 0.036, and the clinically accessible measures by 0.011 to 0.028 (**Figure 4D**). Complete geographic replication estimates are provided in **Supplemental Table 7**.

At the 20% prioritization level, the clinically accessible measures increased the proportion of future events captured across all 4 outcomes (**Figure 4E**), and positive increases were also observed across prioritization levels from 10% to 30% (**Supplemental Figures 6A-6D**). After excluding events occurring within the first 2 or 5 years, incremental discrimination remained evident for CHD, HF, and stroke (**Figure 4F**). AF was not included in this analysis because its primary prediction horizon was 5 years.

## DISCUSSION

In this study of 266,410 adults without prevalent CHD, HF, AF, or stroke, contemporary disease-specific risk equations concentrated 44.8% to 55.2% of subsequent events within the highest-risk 20%. Broad baseline phenotyping provided additional prognostic information beyond these equations, and part of this information could be compressed from 3,838 baseline phenotypes to disease-specific sets of 13 to 19 clinically accessible measures. These clinically accessible measures retained part of the broader phenotypic increment, with the largest discrimination improvement for HF, remained informative after accounting for polygenic risk, and showed directionally consistent performance across UK Biobank regions. Their value in routine clinical practice remains untested (**Central Illustration**).

These findings should be viewed as an extension of established risk assessment rather than an alternative to it. PREVENT, CHARGE-AF, and other clinical equations are intentionally parsimonious and remain central to cardiovascular prevention.^4–8^ The finding that the highest-risk 20% accounted for 44.8% to 55.2% of future events reflects effective risk concentration while also showing that many future cases have less prominent conventional risk profiles. Because CHD, HF, AF, and stroke do not share a common treatment threshold, we used the highest-risk 20% as a common rank-based reference rather than a clinical decision threshold. The relevant clinical question is therefore whether additional, readily obtainable information can refine risk stratification beyond established equations.

The additional information was distributed across several types of baseline measurement. Routine laboratory measures contributed consistently, while questionnaire-derived health information, symptoms, physical and functional measures, physiologic assessments, and selected molecular measurements also contributed. This is consistent with prior studies showing incremental cardiovascular risk information from genetic susceptibility, social and behavioral factors, circulating biomarkers, and electrocardiographic phenotypes.^9–27^ Unlike studies centered on a single outcome or measurement modality, this analysis evaluated the same baseline phenotype framework across 4 cardiovascular outcomes. Incomplete overlap and only partial correlation between contributing components suggest that these measures capture partly distinct dimensions of cardiovascular risk.

Sets of 13 to 19 clinically accessible measures retained part of the broader-model information, with substantial differences across outcomes. Retention of the broader-model discrimination gain was greater for stroke and HF than for AF and CHD. Farah et al^20^ showed that a small set of genetic, lipid, and inflammatory biomarkers can provide complementary information for coronary risk prediction. Our study addresses a complementary question: how much of the information identified across a broad baseline phenotype space can be retained when assessment is restricted to a small number of practical measures. The variation in information retention argues against assuming that one set of measures will perform similarly across cardiovascular outcomes.

The clinically accessible measures also provided information that was largely complementary to inherited genetic susceptibility. The relative contribution of genetic risk varied across cardiovascular outcomes, whereas phenotypic and genetic information remained broadly complementary overall. Measure sets derived with and without PRS adjustment were highly similar and achieved nearly identical discrimination when evaluated in addition to the clinical equation and PRS. In this cohort, separate measure selection after PRS adjustment produced little additional improvement.

The improvement in discrimination was accompanied by a greater proportion of future events captured within the prioritized group. At the 20% prioritization level, the phenotype models captured a greater proportion of future events than the clinical equations, including after restriction to the clinically accessible measures. This comparison complements AUC by showing how changes in risk stratification alter the proportion of future events represented within an equally sized prioritized group. However, improved event capture does not by itself establish clinical benefit. Whether these measures improve prevention will depend on their performance at actionable thresholds and whether the additional information changes management in a way that improves outcomes. Prospective clinical evaluation is therefore required before implementation.

Internal reproducibility analyses showed that high-contribution associations more often replicated across data partitions, recurred across cardiovascular outcomes, and were supported by complementary epidemiologic or genetic analyses. High-contribution associations identified in validation data were also substantially more likely to remain positively informative in the held-out sample. Together, these analyses indicate that high-contribution associations tended to recur across outcomes and remain informative in the held-out sample. Nevertheless, prognostic value should not be interpreted as evidence that the selected phenotypes are causal or modifiable; symptoms, functional limitations, and laboratory abnormalities may also reflect accumulated exposures, comorbidity, or subclinical disease.

Geographic replication provided a further test of reproducibility. Models developed in England retained positive discrimination and increases in the proportion of future events captured at the 20% prioritization level when evaluated without refitting in Scotland and Wales, with similar direction across prioritization levels from 10% to 30%. Because these populations remained within UK Biobank and shared similar recruitment and outcome-ascertainment procedures, these findings represent geographic replication rather than independent external validation. Evaluation across other populations, health care systems, and clinical settings remains necessary.^7,8,40^

## STUDY LIMITATIONS

Several limitations merit emphasis. UK Biobank participants are generally healthier than the general population, and the analysis was restricted to genetically inferred White British participants, limiting generalizability.^41^ Development and geographic replication samples came from the same biobank and therefore do not constitute independent external validation. Published equations required study-specific predictor mapping, and the assessment levels represented prespecified population fractions rather than treatment thresholds. Some baseline phenotypes may reflect unrecognized disease despite the landmark analyses. The clinically accessible measures were selected and evaluated in a research cohort and may perform differently in routine care. Finally, improvements in discrimination and the proportion of future events captured do not establish net clinical benefit, causality, or benefit from modifying any selected phenotype.

## CONCLUSIONS

Recruitment-baseline phenotypes refined cardiovascular risk stratification for CHD, HF, AF, and stroke beyond contemporary clinical equations. Disease-specific sets of clinically accessible measures retained part of this information and showed geographic reproducibility within UK Biobank. Independent external validation and prospective evaluation of calibration, feasibility, and clinical utility are required before use in routine care.

## PERSPECTIVES

## COMPETENCY IN MEDICAL KNOWLEDGE

Contemporary cardiovascular risk equations effectively concentrate future events among individuals at higher predicted risk, yet risk remains distributed beyond the highest-risk 20%. Routinely recorded laboratory measures and brief assessments of symptoms, general health, and physical function provide complementary prognostic information and can refine cardiovascular risk stratification when added to established clinical equations.

## TRANSLATIONAL OUTLOOK

Prospective studies in independent and diverse populations are needed to determine whether adding these clinically accessible measures to established risk equations improves risk-guided preventive decisions and patient outcomes, and whether their incremental value justifies their use in routine clinical care.

## Supporting information

Data Set S1 contains the machine-readable registry of 3,838 baseline phenotypes and is supplied separately

## Data Availability

The data underlying this study were obtained from UK Biobank under Application Number 83974. These data are available to eligible researchers upon application to and approval by UK Biobank (https://www.ukbiobank.ac.uk/). Restrictions apply to their availability, and the individual-level data cannot be publicly shared by the authors. S

## AUTHOR CONTRIBUTIONS

D.H. and Y.Y. conceived and designed the study. H.Y. performed the primary statistical analyses, interpreted the results, and drafted the manuscript. J.G., X.S., K.M., Y.J., and D.L. contributed to data acquisition, statistical analyses, and methodological development. X.Z. contributed to interpretation of the study findings and critically revised the manuscript for important intellectual content. D.H. supervised the study, contributed to data interpretation and methodological development, critically revised the manuscript, and obtained funding. Y.Y. contributed to study supervision, interpretation of the results, and critical revision of the manuscript. All authors reviewed and approved the final manuscript.

## ACKNOWLEDGMENTS

We thank the participants and investigators of UK Biobank (Application 83974). We also acknowledge the investigators of FinnGen and the Million Veteran Program for publicly available genetic summary data used in genetic triangulation.

## ABBREVIATIONS AND ACRONYMS

AF: atrial fibrillation
AUC: area under the receiver operating characteristic curve
CHD: coronary heart disease
CVD: cardiovascular disease
HF: heart failure
IPCW: inverse probability of censoring weighting
LDSC: linkage disequilibrium score regression
MR: Mendelian randomization
PREVENT: Predicting Risk of Cardiovascular Disease Events
PRS: polygenic risk score

---

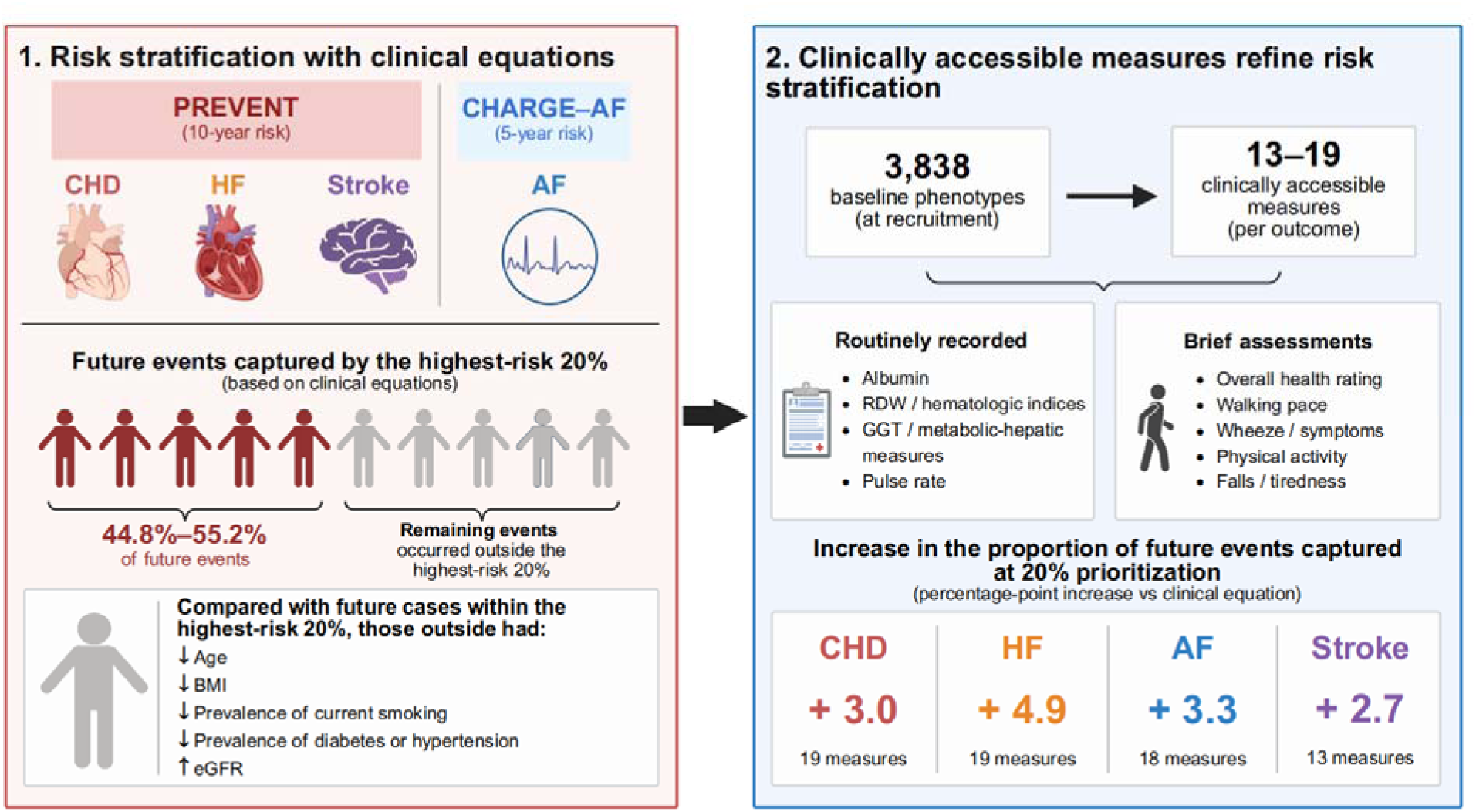

**Central Illustration. Clinically Accessible Measures Refine Cardiovascular Risk Stratification** Clinical equations concentrated 44.8%–55.2% of future events within the highest-risk 20%. Arrows indicate lower or higher characteristics among future cases outside versus within this group. Screening 3,838 baseline phenotypes yielded 13–19 clinically accessible measures per outcome. Adding these measures increased event capture at 20% prioritization by 3.0, 4.9, 3.3, and 2.7 percentage points for CHD, HF, AF, and stroke, respectively. Independent external validation is required before clinical use. AF = atrial fibrillation; BMI = body mass index; CHARGE-AF = Cohorts for Heart and Aging Research in Genomic Epidemiology–Atrial Fibrillation; CHD = coronary heart disease; eGFR = estimated glomerular filtration rate; GGT = gamma-glutamyl transferase; HF = heart failure; PREVENT = Predicting Risk of Cardiovascular Disease Events; RDW = red cell distribution width.

