## Supplemental Methods, Supplemental Tables, Supplemental Figures for "Clinically Accessible Phenotypes Refine Cardiovascular Risk Stratification Beyond Contemporary Clinical Equations"

Data Set S1 contains the machine-readable registry of 3,838 baseline phenotypes and is supplied separately.

### Supplemental Methods

#### Study sample and outcomes

UK Biobank recruited adults aged 40 to 69 years across England, Scotland, and Wales from 2006 through 2010.^1^ We included 266,410 unrelated participants of genetically inferred White British ancestry with quality-controlled genotype data and no prevalent coronary heart disease (CHD), heart failure (HF), atrial fibrillation (AF), or stroke. Participants were assigned once to training, validation, and held-out samples in a 50:20:30 ratio. Model fitting used training data, model selection used validation data, and held-out participants were used for evaluation.

Incident outcomes were identified from linked hospital admission and mortality records through December 19, 2022. CHD was defined as PREVENT-compatible myocardial infarction or fatal CHD, HF as incident HF, AF as incident AF, and stroke as PREVENT-compatible fatal or nonfatal stroke. The horizon was 10 years for CHD, HF, and stroke and 5 years for AF. Participants with a prevalent outcome, inadequate follow-up, or inputs outside the prespecified equation eligibility rules were excluded from that outcome-specific analysis (**Supplemental** **Table 3**; **Supplemental Figure 1**).

#### Clinical equations and polygenic risk scores

The primary comparator was the published disease-specific clinical equation: PREVENT-CHD, the PREVENT-HF base equation without optional predictors, PREVENT-stroke, or CHARGE-AF.^2-4^ Published coefficients were used without outcome-specific refitting. PREVENT-HF was calculated with preventr version 0.12.0. CHARGE-AF used the available UK Biobank mapping; prior myocardial infarction and HF were set to 0 because the cohort was free of these diseases at baseline (**Supplemental Table 1**). Recalibration estimated an intercept and slope in the training sample and was confined to the analysis shown in **Supplemental Figure 2**. We evaluated the incremental phenotypic information beyond the clinical equation and further examined whether this information remained informative after accounting for the disease-specific PRS, which were generated using PRS-CS.^5^ Two PRS analyses were distinguished: independent selection of a PRS-adjusted measure set, and addition of PRS while retaining the Clinical-derived measure set. The four-model conditional comparisons used the same Clinical-derived measures with and without PRS; the separately selected PRS-adjusted sets were used for selection-overlap and cross-application analyses.

#### Baseline phenotype registry

We analyzed 3,838 baseline phenotypes meeting the timing and data-quality criteria. These comprised recruitment measures, variables derived from baseline information, and assays of baseline biospecimens performed later. Repeat assessments, post-recruitment variables, administrative duplicates, and variables without an analyzable baseline representation were excluded before outcome modeling. The retained phenotypes were classified into 6 phenotypic domains (B1-B6) and 6 measurement modalities (M1-M6) (**Supplemental Table 2**). All phenotypic domains and measurement modalities were considered in the broader phenotype models, whereas clinical accessibility was considered only when deriving the smaller disease-specific measure sets.

Continuous and binary variables were processed in training data, and the resulting coding, scaling, and missingness parameters were applied unchanged to validation and evaluation samples. Indicator terms retained information on missingness where specified in the prespecified analysis matrices. No preprocessing step used outcome information from validation or held-out participants. The full phenotype registry, including field identifiers, baseline source details, taxonomy, and clinical availability, is provided in **Data Set S1**.

#### Phenotype model development

Within each outcome and B-by-M component, we fitted Cox models with the clinical-equation linear predictor as an offset. Candidate sets of 5, 10, and 20 phenotypes were compared in validation data. The selected component used the smallest set retaining at least 90% of the best positive validation increment in inverse probability of censoring weighted (IPCW) area under the receiver operating characteristic curve (AUC). Component features were then combined in an elastic-net Cox model.^6^ The model architecture was selected in validation data, coefficients were re-estimated in the combined training and validation sample, and performance was evaluated in held-out participants.

#### Assessment levels and component attribution

Risk stratification was compared at validation-derived prioritization levels of 10%, 15%, 20%, 25%, and 30%; 20% was the principal reporting reference. Validation-sample cutoffs were applied unchanged to held-out participants. An event was newly included if its participant was below the clinical-equation cutoff but above the phenotype-model cutoff, and displaced under the reverse pattern. The increase in the proportion of future events captured was 100 times the difference between newly included and displaced events divided by all observed horizon events. Realized prioritized proportions could differ between models because evaluation-sample thresholds were not refitted to force equal group sizes. The full 10%-30% range was used for sensitivity analyses. The normalized trapezoidal mean across this range remained part of the original development-stage selection rule.

Exact group Shapley values partitioned the change in future events captured at 20% prioritization across phenotypic domain-by-modality components. Contributions were expressed in percentage points and summed to the corresponding broader-model increment. For the component-overlap analysis, a future case could be newly included by more than one component; the union and multiplicity of these cases are not the net event gain of the joint model.

#### Supporting evidence

High-contribution phenotype-outcome associations were compared with low- or no-contribution associations for Bonferroni association, split-sample replication, cross-outcome recurrence, linkage disequilibrium score regression support, and Mendelian randomization support.^7-11^ Evidence layers were evaluated after predictive model development and did not enter feature selection. Genetic analyses used external summary statistics and standard harmonization procedures. These analyses assess convergence and reproducibility, not causality.

#### Risk refinement with clinically accessible measures

We sought a smaller set of measures that retained useful incremental discrimination while remaining clinically accessible. Candidates were restricted to routinely recorded measures and brief additional assessments, including brief self-reported, symptom, or functional assessments and selected additional blood or urine assays. Specialized assays, imaging, and predictors duplicating the clinical equation were excluded. Training-sample elastic-net Cox paths used the clinical-equation linear predictor as an offset. Validation selection required no more than 20 measures and a positive average increase in event capture across 10%-30% prioritization. The retention threshold was the larger of 90% of the clinically accessible reference model's validation AUC increment and that increment minus 0.001. Among eligible sets, the smallest was selected, with validation AUC and event capture used to resolve ties. If no set met the target, candidates within 0.001 of the best capped validation AUC were retained; among these, candidates within 0.10 percentage points of the best average event-capture increment were retained, and the smallest set was selected. Final sets contained 19 CHD, 19 HF, 18 AF, and 13 stroke measures.

#### Geographic replication

Model development was repeated in England. England training and validation samples were used for phenotype selection, tuning, coefficient estimation, measure-set size, and prioritization thresholds. The resulting models were then evaluated once in Scotland and Wales without refitting or recalibration. This analysis assesses geographic transportability and internal replication within UK Biobank; it is not external validation in an independent health care system or population.

#### Statistical analysis

Discrimination was summarized with horizon-specific IPCW AUC and Harrell C-index; calibration was summarized with calibration slope and the observed-to-expected ratio.^12-14^ Differences between models used 1,000 paired participant-level bootstrap samples. IPCW discrimination analyses re-estimated censoring weights in each replicate. For event capture at 20%, model scores and validation-derived thresholds remained fixed, and the denominator was the number of observed horizon events in each resample. Percentile 95% confidence intervals are reported. Tests were 2-sided. Analyses were performed in R.

### Supplemental Tables

#### Supplemental Table 1. Published predictors in the clinical equations

| **Published predictor** | **PREVENT-CHD** | **PREVENT-HF** | **CHARGE-AF** | **PREVENT-stroke** |
| --- | --- | --- | --- | --- |
| Age | Included | Included | Included | Included |
| Sex | Included | Included | Not included | Included |
| Total cholesterol | Derivation input | Not included | Not included | Derivation input |
| Non-HDL cholesterol | Included | Not included | Not included | Included |
| HDL cholesterol | Included | Not included | Not included | Included |
| Systolic blood pressure | Included | Included | Included | Included |
| Diabetes | Included | Included | Included | Included |
| Current smoking | Included | Included | Included | Included |
| eGFR | Included | Included | Not included | Included |
| Antihypertensive treatment | Included | Included | Included | Included |
| Statin use | Included | Not included | Not included | Included |
| Body mass index | Applicability screening only | Included | Not included | Applicability screening only |
| White race | Not included | Not included | Included; project proxy mapping | Not included |
| Height | Not included | Not included | Included | Not included |
| Weight | Not included | Not included | Included | Not included |
| Diastolic blood pressure | Not included | Not included | Included | Not included |
| Prior heart failure | Not included | Not included | Published term; fixed to 0 | Not included |
| Prior myocardial infarction | Not included | Not included | Published term; fixed to 0 | Not included |

PREVENT-HF used the base equation without optional predictors. Non-HDL cholesterol was derived from total and HDL cholesterol. The project CHARGE-AF implementation used a binary White-ethnicity proxy; prior HF and myocardial infarction were fixed to 0 in the baseline CVD-free cohort. BMI indicates body mass index; eGFR, estimated glomerular filtration rate; HDL, high-density lipoprotein.

#### Supplemental Table 2. Baseline phenotype taxonomy

| **Taxonomy** | **Code** | **Definition** | **Phenotypes, n** |
| --- | --- | --- | --- |
| Phenotypic domain | B1 | Behavioral/lifestyle | 154 |
| Phenotypic domain | B2 | Metabolic/renal/hematologic | 455 |
| Phenotypic domain | B3 | Cardiopulmonary/functional reserve | 180 |
| Phenotypic domain | B4 | Symptoms/global health | 30 |
| Phenotypic domain | B5 | Social/life-course | 203 |
| Phenotypic domain | B6 | Other/system-specific biology | 2,816 |
| Measurement modality | M1 | Questionnaire/self-report | 381 |
| Measurement modality | M2 | Routine laboratory | 59 |
| Measurement modality | M3 | Routine examination | 213 |
| Measurement modality | M4 | Simple physiology/function | 21 |
| Measurement modality | M5 | Proteomics | 2,917 |
| Measurement modality | M6 | Metabolomics | 247 |

Counts are marginal; each of the 3,838 retained phenotypes contributes once to a domain and once to a modality. Data Set S1 provides the complete machine-readable registry and B-by-M assignment.

#### Supplemental Table 3. Analysis samples, endpoints, and clinical comparators

| **Analysis** | **Outcome** | **Endpoint** | **Comparator** | **Horizon, y** | **Participants, n** | **Events, n** |
| --- | --- | --- | --- | --- | --- | --- |
| Primary held-out | CHD | PREVENT-compatible myocardial infarction or fatal CHD | PREVENT-CHD | 10 | 75,089 | 1,286 |
| Primary held-out | HF | Incident HF | PREVENT-HF base equation | 10 | 77,900 | 1,022 |
| Primary held-out | AF | Incident AF | CHARGE-AF | 5 | 75,367 | 1,105 |
| Primary held-out | Stroke | PREVENT-compatible fatal or nonfatal stroke | PREVENT-stroke | 10 | 75,089 | 961 |
| Scotland/Wales | CHD | Same as primary | PREVENT-CHD | 10 | 30,119 | 551 |
| Scotland/Wales | HF | Same as primary | PREVENT-HF base equation | 10 | 31,195 | 326 |
| Scotland/Wales | AF | Same as primary | CHARGE-AF | 5 | 21,692 | 293 |
| Scotland/Wales | Stroke | Same as primary | PREVENT-stroke | 10 | 30,119 | 491 |

The base cohort was assigned to training (n=133,205), validation (n=53,282), and held-out evaluation (n=79,923) in a 50:20:30 ratio. Outcome-specific counts reflect equation eligibility and endpoint availability. England development used training and validation samples; Scotland and Wales were evaluated once without refitting. AF indicates atrial fibrillation; CHD, coronary heart disease; HF, heart failure.

#### Supplemental Table 4. Final clinically accessible measure sets

| **Outcome** | **Order** | **Measure** | **UKB field** | **Modality** | **Acquisition tier** | **Clinical dimension** |
| --- | --- | --- | --- | --- | --- | --- |
| CHD | 1 | Albumin | 30600 | Routine laboratory | Routinely recorded | Metabolic/hepatic |
| CHD | 2 | Alkaline phosphatase | 30610 | Routine laboratory | Routinely recorded | Metabolic/hepatic |
| CHD | 3 | Haematocrit percentage | 30030 | Routine laboratory | Routinely recorded | Hematologic/inflammatory |
| CHD | 4 | Haemoglobin concentration | 30020 | Routine laboratory | Routinely recorded | Hematologic/inflammatory |
| CHD | 5 | Monocyte count | 30130 | Routine laboratory | Routinely recorded | Hematologic/inflammatory |
| CHD | 6 | Neutrophil count | 30140 | Routine laboratory | Routinely recorded | Hematologic/inflammatory |
| CHD | 7 | Red blood cell (erythrocyte) distribution width | 30070 | Routine laboratory | Routinely recorded | Hematologic/inflammatory |
| CHD | 8 | Alcohol intake frequency | 1558 | Questionnaire | Brief assessment | Functional/behavioral |
| CHD | 9 | Cystatin C | 30720 | Routine laboratory | Brief assessment | Cardiorenal |
| CHD | 10 | Duration of moderate activity | 894 | Questionnaire | Brief assessment | Functional/behavioral |
| CHD | 11 | Duration of walks | 874 | Questionnaire | Brief assessment | Functional/behavioral |
| CHD | 12 | Frequency of tiredness / lethargy in last 2 weeks | 2080 | Questionnaire | Brief assessment | Functional/behavioral |
| CHD | 13 | Long-standing illness, disability or infirmity | 2188 | Questionnaire | Brief assessment | Functional/behavioral |
| CHD | 14 | Mean reticulocyte volume | 30260 | Routine laboratory | Brief assessment | Hematologic/inflammatory |
| CHD | 15 | Microalbumin in urine | 30500 | Routine laboratory | Brief assessment | Cardiorenal |
| CHD | 16 | Overall health rating | 2178 | Questionnaire | Brief assessment | Functional/behavioral |
| CHD | 17 | Urate | 30880 | Routine laboratory | Brief assessment | Cardiorenal |
| CHD | 18 | Usual walking pace | 924 | Simple physiology/function | Brief assessment | Functional/behavioral |
| CHD | 19 | Wheeze or whistling in the chest in last year | 2316 | Questionnaire | Brief assessment | Cardiopulmonary |
| HF | 1 | Albumin | 30600 | Routine laboratory | Routinely recorded | Metabolic/hepatic |
| HF | 2 | Alkaline phosphatase | 30610 | Routine laboratory | Routinely recorded | Metabolic/hepatic |
| HF | 3 | Gamma glutamyltransferase | 30730 | Routine laboratory | Routinely recorded | Metabolic/hepatic |
| HF | 4 | Lymphocyte percentage | 30180 | Routine laboratory | Routinely recorded | Hematologic/inflammatory |
| HF | 5 | Mean corpuscular volume | 30040 | Routine laboratory | Routinely recorded | Hematologic/inflammatory |
| HF | 6 | Neutrophil count | 30140 | Routine laboratory | Routinely recorded | Hematologic/inflammatory |
| HF | 7 | Pulse rate, automated reading | 102 | Routine examination | Routinely recorded | Cardiopulmonary |
| HF | 8 | Red blood cell (erythrocyte) distribution width | 30070 | Routine laboratory | Routinely recorded | Hematologic/inflammatory |
| HF | 9 | Duration of vigorous activity | 914 | Questionnaire | Brief assessment | Functional/behavioral |
| HF | 10 | Falls in the last year | 2296 | Questionnaire | Brief assessment | Functional/behavioral |
| HF | 11 | Former alcohol drinker | 3731 | Questionnaire | Brief assessment | Functional/behavioral |
| HF | 12 | Long-standing illness, disability or infirmity | 2188 | Questionnaire | Brief assessment | Functional/behavioral |
| HF | 13 | Mean reticulocyte volume | 30260 | Routine laboratory | Brief assessment | Hematologic/inflammatory |
| HF | 14 | Microalbumin in urine | 30500 | Routine laboratory | Brief assessment | Cardiorenal |
| HF | 15 | Overall health rating | 2178 | Questionnaire | Brief assessment | Functional/behavioral |
| HF | 16 | Phosphate | 30810 | Routine laboratory | Brief assessment | Cardiorenal |
| HF | 17 | Urate | 30880 | Routine laboratory | Brief assessment | Cardiorenal |
| HF | 18 | Usual walking pace | 924 | Simple physiology/function | Brief assessment | Functional/behavioral |
| HF | 19 | Wheeze or whistling in the chest in last year | 2316 | Questionnaire | Brief assessment | Cardiopulmonary |
| AF | 1 | Albumin | 30600 | Routine laboratory | Routinely recorded | Metabolic/hepatic |
| AF | 2 | Gamma glutamyltransferase | 30730 | Routine laboratory | Routinely recorded | Metabolic/hepatic |
| AF | 3 | Lymphocyte percentage | 30180 | Routine laboratory | Routinely recorded | Hematologic/inflammatory |
| AF | 4 | Mean corpuscular volume | 30040 | Routine laboratory | Routinely recorded | Hematologic/inflammatory |
| AF | 5 | Monocyte count | 30130 | Routine laboratory | Routinely recorded | Hematologic/inflammatory |
| AF | 6 | Platelet count | 30080 | Routine laboratory | Routinely recorded | Hematologic/inflammatory |
| AF | 7 | Pulse rate, automated reading | 102 | Routine examination | Routinely recorded | Cardiopulmonary |
| AF | 8 | Red blood cell (erythrocyte) distribution width | 30070 | Routine laboratory | Routinely recorded | Hematologic/inflammatory |
| AF | 9 | Total bilirubin | 30840 | Routine laboratory | Routinely recorded | Metabolic/hepatic |
| AF | 10 | Triglycerides | 30870 | Routine laboratory | Routinely recorded | Metabolic/hepatic |
| AF | 11 | Cystatin C | 30720 | Routine laboratory | Brief assessment | Cardiorenal |
| AF | 12 | Long-standing illness, disability or infirmity | 2188 | Questionnaire | Brief assessment | Functional/behavioral |
| AF | 13 | Mean reticulocyte volume | 30260 | Routine laboratory | Brief assessment | Hematologic/inflammatory |
| AF | 14 | Microalbumin in urine | 30500 | Routine laboratory | Brief assessment | Cardiorenal |
| AF | 15 | Overall health rating | 2178 | Questionnaire | Brief assessment | Functional/behavioral |
| AF | 16 | Summed days activity | 22033 | Questionnaire | Brief assessment | Functional/behavioral |
| AF | 17 | Urate | 30880 | Routine laboratory | Brief assessment | Cardiorenal |
| AF | 18 | Wheeze or whistling in the chest in last year | 2316 | Questionnaire | Brief assessment | Cardiopulmonary |
| Stroke | 1 | Albumin | 30600 | Routine laboratory | Routinely recorded | Metabolic/hepatic |
| Stroke | 2 | Gamma glutamyltransferase | 30730 | Routine laboratory | Routinely recorded | Metabolic/hepatic |
| Stroke | 3 | Mean corpuscular volume | 30040 | Routine laboratory | Routinely recorded | Hematologic/inflammatory |
| Stroke | 4 | Monocyte count | 30130 | Routine laboratory | Routinely recorded | Hematologic/inflammatory |
| Stroke | 5 | Red blood cell (erythrocyte) distribution width | 30070 | Routine laboratory | Routinely recorded | Hematologic/inflammatory |
| Stroke | 6 | Cystatin C | 30720 | Routine laboratory | Brief assessment | Cardiorenal |
| Stroke | 7 | Daytime dozing / sleeping | 1220 | Questionnaire | Brief assessment | Functional/behavioral |
| Stroke | 8 | Falls in the last year | 2296 | Questionnaire | Brief assessment | Functional/behavioral |
| Stroke | 9 | Former alcohol drinker | 3731 | Questionnaire | Brief assessment | Functional/behavioral |
| Stroke | 10 | Long-standing illness, disability or infirmity | 2188 | Questionnaire | Brief assessment | Functional/behavioral |
| Stroke | 11 | Microalbumin in urine | 30500 | Routine laboratory | Brief assessment | Cardiorenal |
| Stroke | 12 | Overall health rating | 2178 | Questionnaire | Brief assessment | Functional/behavioral |
| Stroke | 13 | Wheeze or whistling in the chest in last year | 2316 | Questionnaire | Brief assessment | Cardiopulmonary |

Order is the final disease-specific measure-set order. Measurement modality follows the frozen M1-M6 taxonomy in Data Set S1. Acquisition tier follows the outcome-blind clinical-admissibility audit used to derive the smaller measure sets. It distinguishes routinely recorded measures from brief assessments in this study. Clinical dimension was assigned after selection for interpretation and did not enter model fitting. UKB indicates UK Biobank.

#### Supplemental Table 5. Held-out performance of the clinical equation, broader phenotype model, and clinically accessible measures

| **Outcome** | **Model** | **Measures, n** | **IPCW AUC** | **Delta AUC vs clinical (95% CI)** | **Change in event capture at 20%, pp (95% CI)** | **Net events at 20%, n** |
| --- | --- | --- | --- | --- | --- | --- |
| CHD | Clinical equation | Published equation | 0.755 | Reference | Reference | Reference |
| CHD | Broader phenotype model | 59 | 0.786 | 0.0305 (0.0236 to 0.0371) | 7.2 (4.8 to 9.6) | 93 |
| CHD | Clinically accessible measures | 19 | 0.768 | 0.0129 (0.0077 to 0.0174) | 3.0 (1.0 to 4.9) | 38 |
| HF | Clinical equation | Published equation | 0.772 | Reference | Reference | Reference |
| HF | Broader phenotype model | 97 | 0.813 | 0.0406 (0.0322 to 0.0490) | 8.0 (5.3 to 11.0) | 82 |
| HF | Clinically accessible measures | 19 | 0.802 | 0.0301 (0.0225 to 0.0381) | 4.9 (2.1 to 7.9) | 50 |
| AF | Clinical equation | Published equation | 0.746 | Reference | Reference | Reference |
| AF | Broader phenotype model | 553 | 0.776 | 0.0295 (0.0236 to 0.0361) | 4.9 (2.9 to 7.1) | 54 |
| AF | Clinically accessible measures | 18 | 0.764 | 0.0176 (0.0128 to 0.0226) | 3.3 (1.5 to 5.4) | 37 |
| Stroke | Clinical equation | Published equation | 0.720 | Reference | Reference | Reference |
| Stroke | Broader phenotype model | 38 | 0.735 | 0.0153 (0.0093 to 0.0222) | 4.0 (1.3 to 6.5) | 38 |
| Stroke | Clinically accessible measures | 13 | 0.734 | 0.0138 (0.0082 to 0.0200) | 2.7 (0.1 to 5.4) | 26 |

Changes in event capture are differences in the percentage of observed horizon events included at the validation-derived 20% prioritization level, relative to the clinical equation. Net events equal newly included minus displaced events. CI indicates confidence interval; IPCW, inverse probability of censoring weighting; pp, percentage points.

#### Supplemental Table 6. Incremental Value of Polygenic and Phenotypic Information

| **Outcome** | **Model** | **Horizon, y** | **Participants, n** | **Events, n** | **IPCW AUC** | **Events captured at 20%, %** |
| --- | --- | --- | --- | --- | --- | --- |
| CHD | Clinical equation | 10 | 75,089 | 1,286 | 0.755 | 49.2 |
| CHD | Clinical + PRS | 10 | 75,089 | 1,286 | 0.782 | 56.0 |
| CHD | Clinical + clinically accessible measures | 10 | 75,089 | 1,286 | 0.768 | 52.2 |
| CHD | Clinical + PRS + Clinical-derived measure set | 10 | 75,089 | 1,286 | 0.794 | 57.1 |
| CHD | Clinical + PRS + PRS-adjusted measure set | 10 | 75,089 | 1,286 | 0.793 | 57.4 |
| HF | Clinical equation | 10 | 77,900 | 1,022 | 0.772 | 55.2 |
| HF | Clinical + PRS | 10 | 77,900 | 1,022 | 0.775 | 55.5 |
| HF | Clinical + clinically accessible measures | 10 | 77,900 | 1,022 | 0.802 | 60.1 |
| HF | Clinical + PRS + Clinical-derived measure set | 10 | 77,900 | 1,022 | 0.803 | 60.7 |
| HF | Clinical + PRS + PRS-adjusted measure set | 10 | 77,900 | 1,022 | 0.803 | 61.0 |
| AF | Clinical equation | 5 | 75,367 | 1,105 | 0.746 | 50.7 |
| AF | Clinical + PRS | 5 | 75,367 | 1,105 | 0.788 | 57.7 |
| AF | Clinical + clinically accessible measures | 5 | 75,367 | 1,105 | 0.764 | 54.0 |
| AF | Clinical + PRS + Clinical-derived measure set | 5 | 75,367 | 1,105 | 0.801 | 61.8 |
| AF | Clinical + PRS + PRS-adjusted measure set | 5 | 75,367 | 1,105 | 0.801 | 61.4 |
| Stroke | Clinical equation | 10 | 75,089 | 961 | 0.720 | 44.8 |
| Stroke | Clinical + PRS | 10 | 75,089 | 961 | 0.724 | 46.0 |
| Stroke | Clinical + clinically accessible measures | 10 | 75,089 | 961 | 0.734 | 47.6 |
| Stroke | Clinical + PRS + Clinical-derived measure set | 10 | 75,089 | 961 | 0.737 | 48.0 |
| Stroke | Clinical + PRS + PRS-adjusted measure set | 10 | 75,089 | 961 | 0.738 | 48.8 |

Event capture is the percentage of observed horizon events included using each model's validation-derived 20% cutoff. The Clinical-derived set is held fixed in the four-model conditional comparisons. The PRS-adjusted set was selected independently after incorporating PRS. PRS indicates polygenic risk score.

#### Supplemental Table 7. Geographic Replication in Scotland and Wales

| **Outcome** | **England-derived model** | **Participants, n** | **Events, n** | **Delta IPCW AUC (95% CI)** | **Change in event capture at 20%, pp (95% CI)** |
| --- | --- | --- | --- | --- | --- |
| CHD | Broader phenotype model | 30,119 | 551 | 0.0334 (0.0234 to 0.0434) | 8.53 (4.87 to 11.90) |
| CHD | Clinically accessible measures | 30,119 | 551 | 0.0144 (0.0069 to 0.0214) | 2.72 (0.00 to 5.51) |
| HF | Broader phenotype model | 31,195 | 326 | 0.0361 (0.0223 to 0.0497) | 6.13 (1.26 to 10.83) |
| HF | Clinically accessible measures | 31,195 | 326 | 0.0279 (0.0143 to 0.0415) | 3.68 (-0.93 to 8.37) |
| AF | Broader phenotype model | 21,692 | 293 | 0.0251 (0.0129 to 0.0371) | 7.51 (3.15 to 11.88) |
| AF | Clinically accessible measures | 21,692 | 293 | 0.0109 (0.0021 to 0.0201) | 3.07 (0.00 to 6.67) |
| Stroke | Broader phenotype model | 30,119 | 491 | 0.0147 (0.0056 to 0.0238) | 6.31 (3.23 to 9.65) |
| Stroke | Clinically accessible measures | 30,119 | 491 | 0.0151 (0.0067 to 0.0238) | 3.26 (0.20 to 6.60) |

Changes in event capture use the validation-derived 20% level. Models were applied without refitting or recalibration. Confidence intervals use 1,000 paired participant-level bootstrap samples.

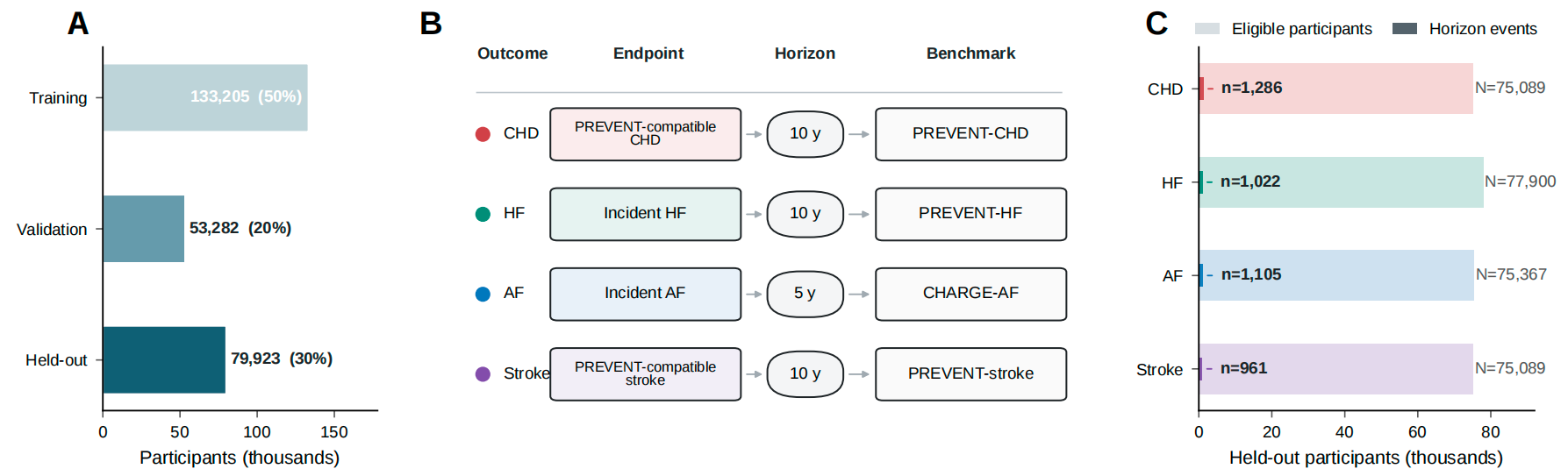

### Supplemental Figures

#### Supplemental Figure 1. Analysis Samples and Outcome-Specific Evaluation Sets

**A**, Allocation of participants to training, validation, and held-out samples in the 50:20:30 split. **B**, Outcome definition, prediction horizon, and clinical comparator for each cardiovascular outcome. **C**, Outcome-specific numbers of eligible held-out participants and horizon events. AF indicates atrial fibrillation; CHD, coronary heart disease; and HF, heart failure.

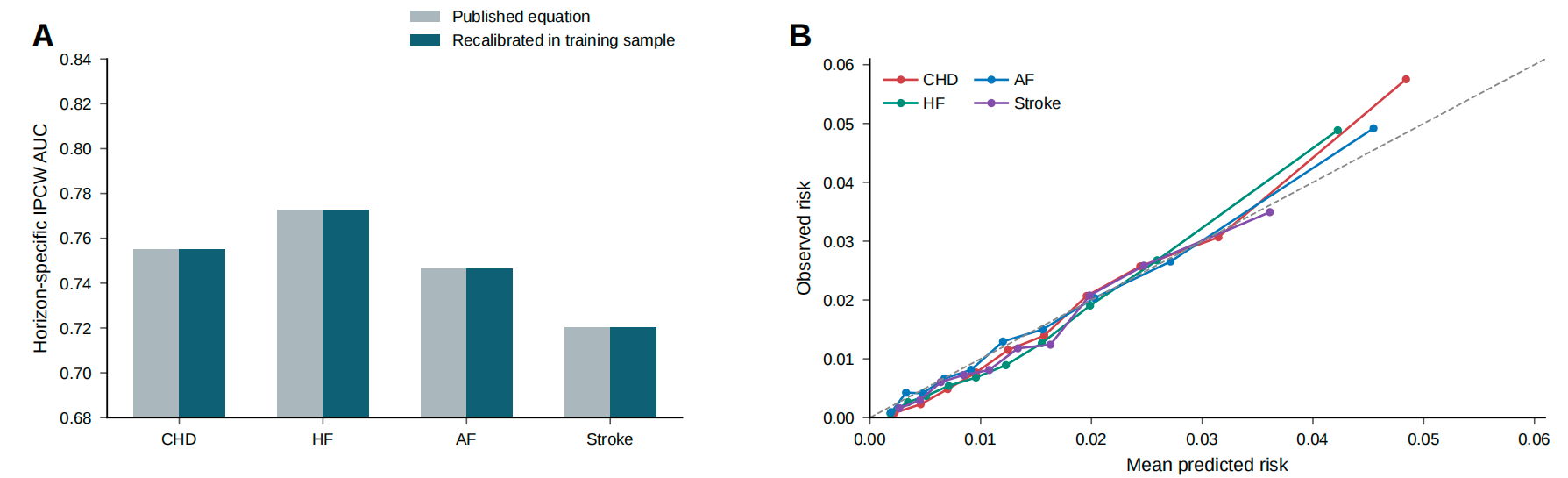

#### Supplemental Figure 2. Discrimination and Calibration of the Clinical Equations

**A**, Held-out horizon-specific IPCW AUC for the published clinical equations and after recalibration in the training sample. **B**, Observed versus mean predicted risk across risk groups; the diagonal line indicates perfect calibration. AUC indicates area under the receiver operating characteristic curve; IPCW, inverse probability of censoring weighting.

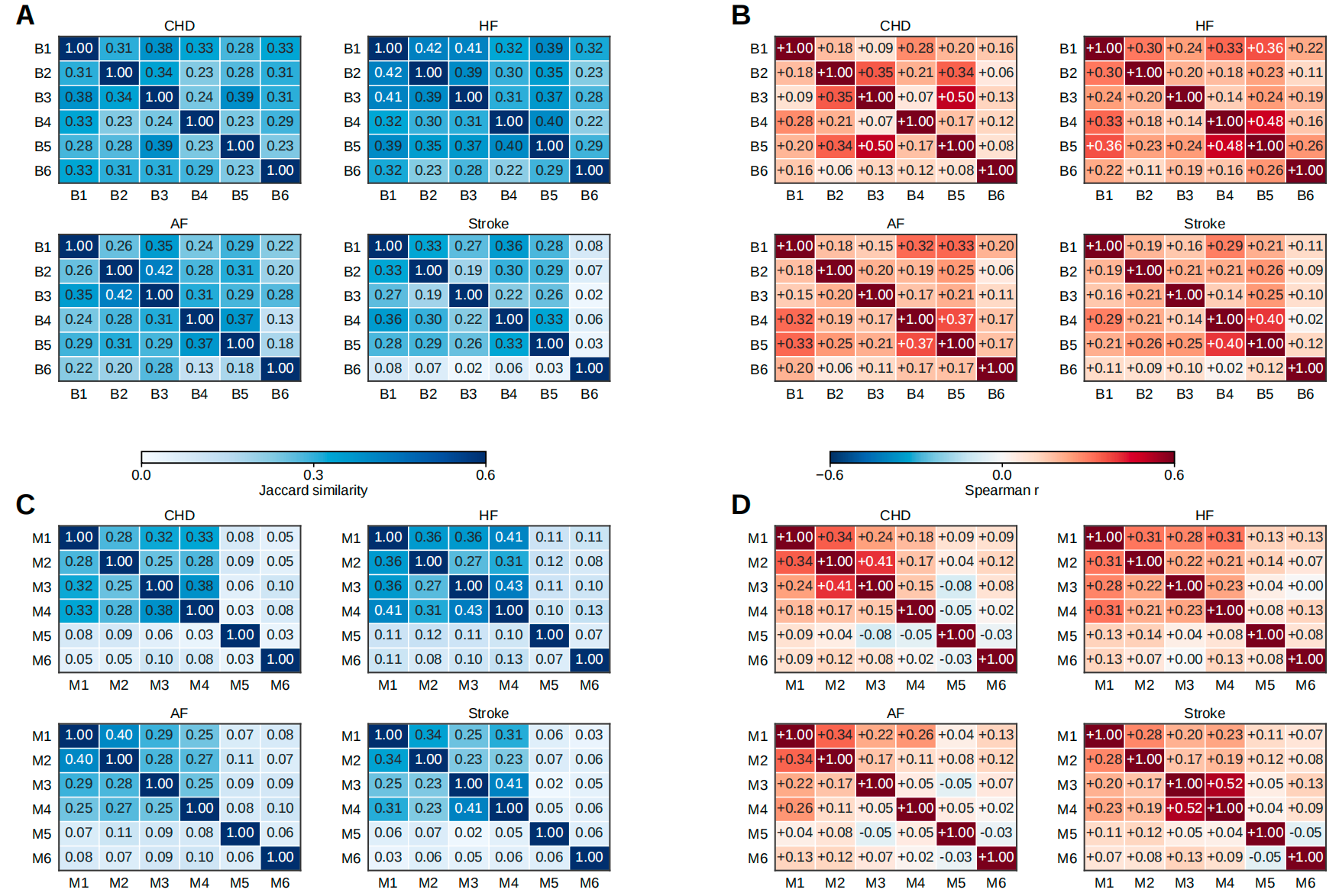

#### Supplemental Figure 3. Overlap and Concordance Across Phenotypic Components

**A and** **B**, Pairwise Jaccard overlap and participant-level score correlation among the 6 phenotypic-domain components, shown separately for CHD, HF, AF, and stroke. **C and D**, Corresponding overlap and score correlations among the 6 measurement-modality components. Analyses were based on held-out predictions from the final models. AF indicates atrial fibrillation; CHD, coronary heart disease; and HF, heart failure.

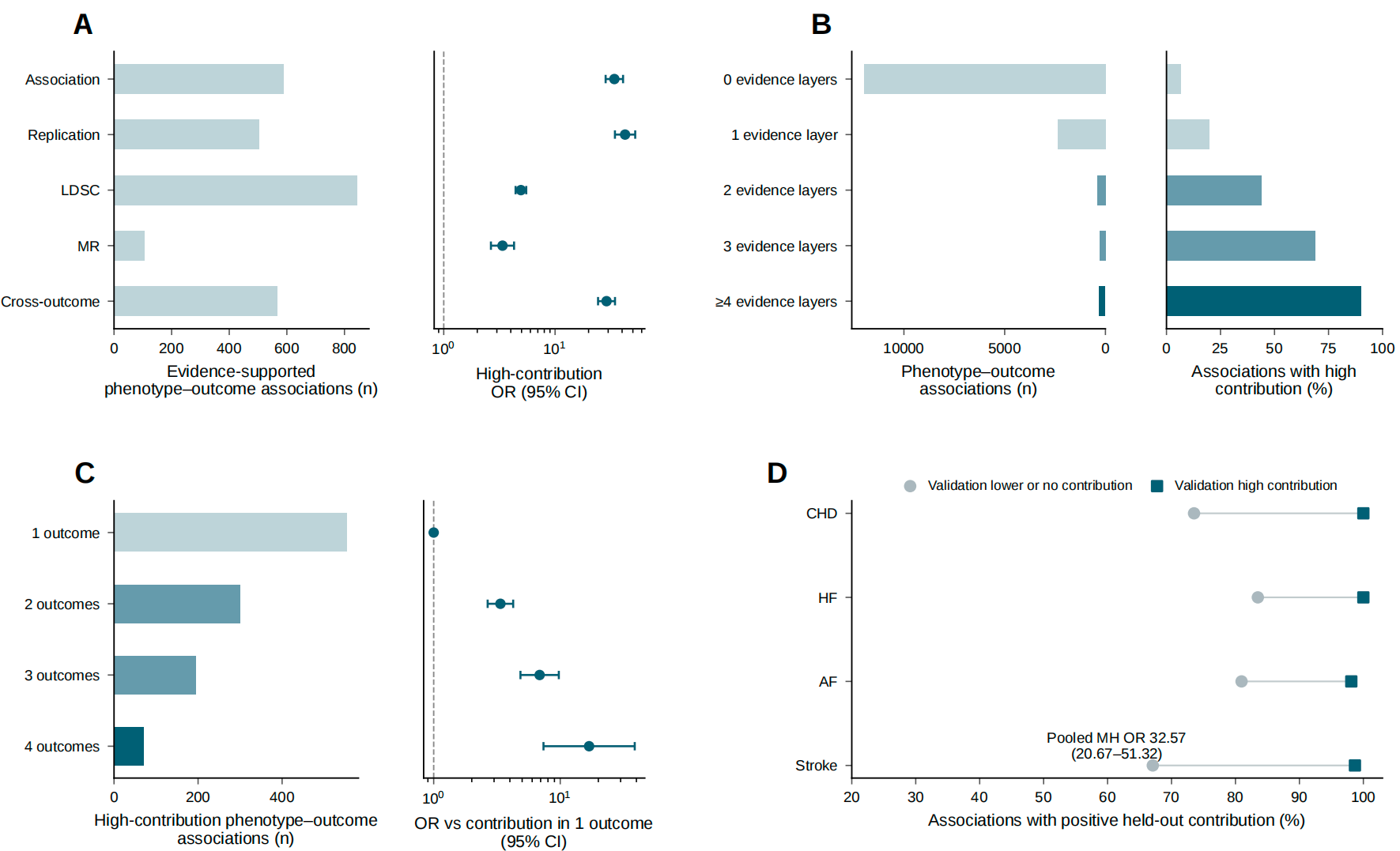

#### Supplemental Figure 4. Convergence and Reproducibility of Phenotype-Outcome Associations

**A**, Number of phenotype-outcome associations meeting each evidence criterion and odds ratios for high contribution among associations with versus without that evidence. **B**, Number of associations and proportion with high contribution according to the number of supporting evidence layers. **C**, Number of high-contribution associations according to recurrence across cardiovascular outcomes and corresponding odds ratios relative to associations occurring in 1 outcome. **D**, Proportion retaining a positive contribution in the held-out sample among associations classified in validation as high contribution versus lower or no contribution; the pooled Mantel-Haenszel odds ratio is shown. Evidence sources were evaluated after model development and did not influence phenotype selection.

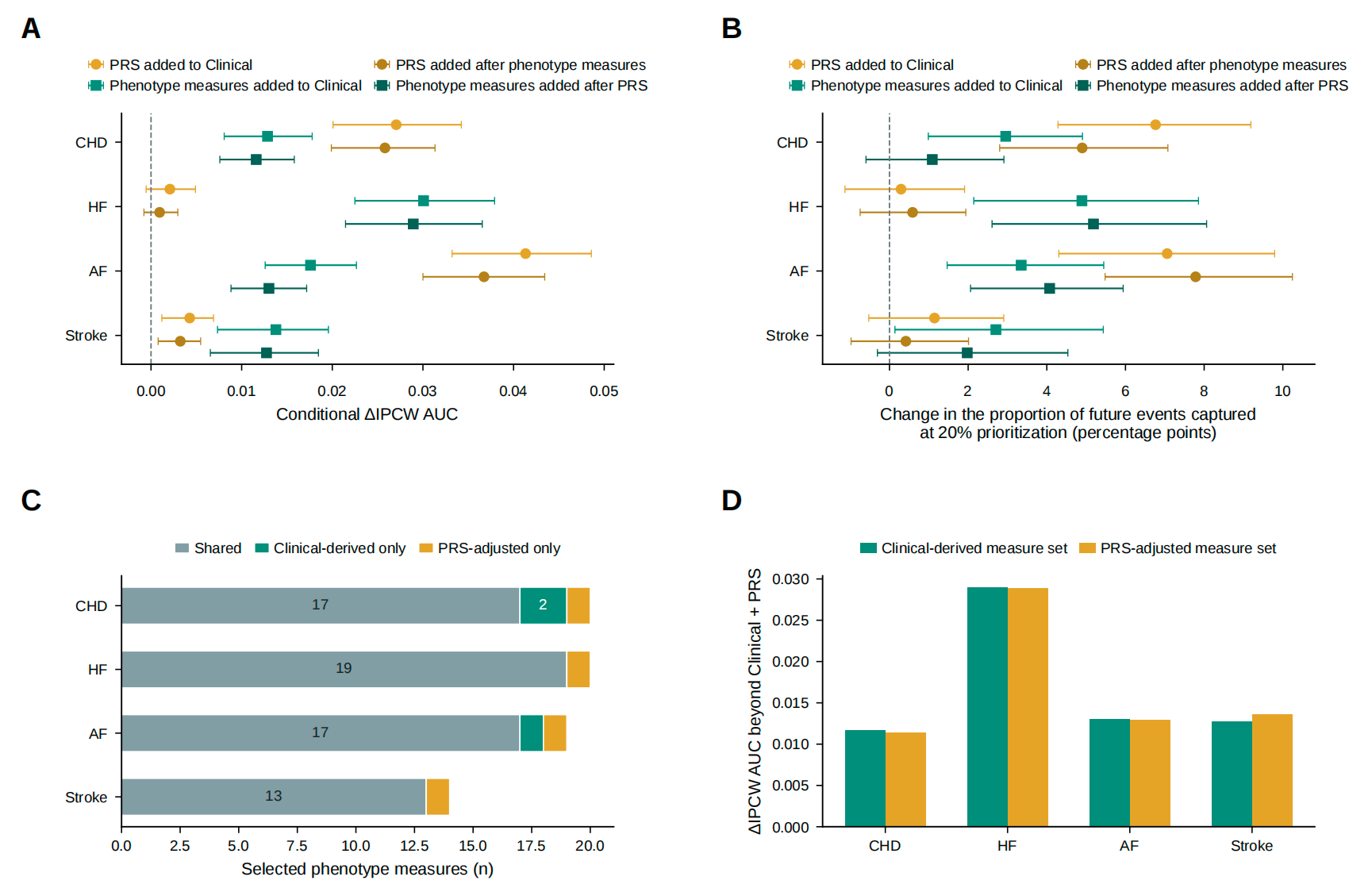

#### Supplemental Figure 5. Complementarity of Polygenic and Phenotypic Information

A, Conditional change in held-out IPCW AUC. B, Conditional change in the proportion of future events captured at 20% prioritization (percentage points). A and B use the same Clinical-derived measures with and without PRS; error bars indicate paired-bootstrap 95% confidence intervals. C, Overlap of Clinical-derived and independently selected PRS-adjusted measure sets. D, IPCW AUC increment beyond Clinical + PRS for these two sets. AF indicates atrial fibrillation; AUC, area under the receiver operating characteristic curve; CHD, coronary heart disease; HF, heart failure; IPCW, inverse probability of censoring weighting; and PRS, polygenic risk score.

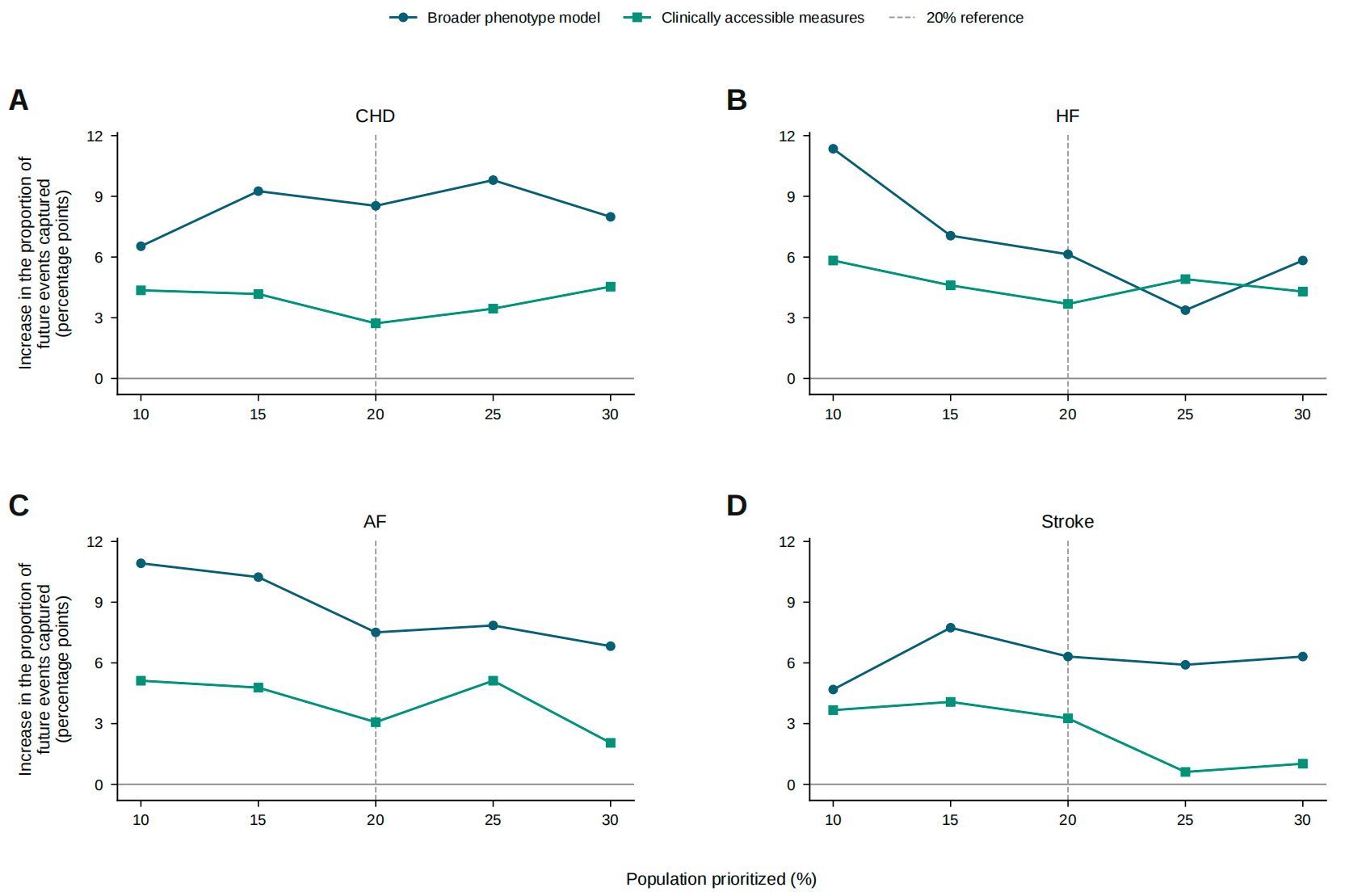

#### Supplemental Figure 6. Sensitivity to Prioritization Level in Scotland and Wales

A to D, Increase in the proportion of future events captured across 10%-30% prioritization levels in Scotland and Wales. Positive values indicate greater capture than the clinical equation at the same validation-derived prioritization level; actual prioritized group sizes can differ. The dashed line marks the 20% reference. Models were applied without refitting or recalibration. CHD indicates coronary heart disease; HF, heart failure; and AF, atrial fibrillation.
